# Cancer screening attendance rates in transgender and gender-diverse patients: a systematic review and meta-analysis

**DOI:** 10.1101/2024.04.17.24305969

**Authors:** Alvina Chan, Charlotte Jamieson, Hannah Draper, Stewart O’Callaghan, Barbara-ann Guinn

## Abstract

**Objectives:** To examine disparities between transgender and gender-diverse (TGD) and cisgender (CG) people through analysis of attendance rates for cancer screening and compare differences between types of cancer screened.

**Design:** Systematic review and meta-analysis.

**Data sources:** PubMed, EMBASE [via Ovid], CINAHL Complete [via EBSCO], and Cochrane Library from inception to 30 September 2023.

**Methods:** Studies for inclusion were case-control or cross-sectional studies with quantitative data investigating TGD adults attending any cancer screening services. Exclusion criteria were studies with participants ineligible for cancer screening or without samples from TGD individuals, qualitative data, and cancer diagnosis from symptomatic presentation or incidental findings. A modified Newcastle-Ottawa Scale was used to assess risk of bias and reports rated poor were excluded. Results were synthesised through random-effects meta-analysis and narrative synthesis.

**Results:** Searches identified 25 eligible records, whereby 18 met risk of bias requirements. These were cross-sectional studies, including retrospective chart reviews and survey analyses, and encompassed over 14.8 million participants. The main outcomes measured were up-to-date (UTD) and lifetime (LT) attendance. Meta-analysis found differences for UTD cervical (OR=0.37, 95% CI [0.23, 0.60], p<0.0001) and mammography screening (OR=0.41, 95% CI [0.20, 0.87], p=0.02). There were no meaningful differences seen in LT results. Pooling total odds ratios for each synthesis (cervical, breast, prostate, and colorectal cancer) showed reduced attendance in TGD participants (OR=0.50, 95% CI [0.37, 0.68], p<0.0001). Narrative synthesis of seven remaining articles supported meta-analysis results, finding generally reduced screening rates in TGD versus CG participants.

**Conclusions:** TGD individuals are overall less likely to utilise cancer screening compared to CG counterparts. The greatest disparity in attendance was seen specifically in UTD cervical screening. Limitations of this review included high risk of bias within studies, high heterogeneity, and a lack of resources for further statistical testing. Individual and structural factors such as psychological distress, socioeconomic status, and healthcare accessibility can prevent TGD people from accessing cancer screening. Bridging this gap will require consolidated efforts from healthcare systems including reviews of structural design, innovation of accessible and inclusive technology, education of HCPs, and reassessment of patient information resources. Joint production of future interventions *with* the TGD community is vital to improving both cancer screening experience and outcomes.

**Funding:** This work was supported by the INSPIRE grant generously awarded to the Hull York Medical School by the Academy of Medical Sciences through the Wellcome Trust [Ref: IR5\1018].

**Systematic review registration:** PROSPERO CRD42022368911.

**KEY MESSAGES:** *What is already known about this topic?:* Many transgender and gender-diverse (TGD) people experience difficulties accessing cancer screening and so face potentially increased risks in morbidity and mortality.

*What this study adds?:* This systematic review and meta-analysis investigated differences in attendance of cancer screening services between TGD and CG people and explored reasons underpinning present disparities. TGD individuals are less likely to attend cancer screening services overall, and are less likely to be up-to-date with breast and cervical cancer screening.

*How this study might affect research, practise of policy?:* To reduce inequities, individual and institutional barriers must be addressed through research, technological innovation, reviews of current structural design, and improved education. It is vital that future interventions for TGD people are jointly produced with the community to improve both cancer screening experience and outcomes.

## INTRODUCTION

An estimated 0.3-0.8% of UK and US people are transgender compared to a worldwide frequency of 0.8–2% [1–3]. Individuals from transgender and gender-diverse (TGD) communities commonly experience inequalities in healthcare. Notably, 23% of TGD individuals in the US stated they avoided seeking necessary medical care in the past year due to discrimination and stigma [4]. This is reflected in cancer screening rates where disparities between TGD and cisgender (CG) individuals are evident. For instance, multiple studies have found TGD adults are less likely to attend cancer screening at recommended intervals [5–11].

Moreover, national cancer screening guidelines that are currently available for TGD people are derived from research on CG participants and are only informed by a limited number of studies specific to TGD populations [12]. Many TGD patients have also encountered negative experiences, like harassment or clinicians displaying inadequate knowledge of their management, leading to decreased trust and utilisation of services [12,13].

Prejudice and discrimination on systemic, structural, and individual levels [14] disproportionately impacts upon the wellbeing of marginalised groups like TGD populations. Disparities in utilisation of cancer screening services may have implications on the morbidity and mortality of TGD individuals [15]. For instance, due to avoiding distress and dysphoria caused by medical procedures involving more “gendered” anatomical structures, TGD people may be at higher risk of breast, cervical, or prostate cancers [16].

It is important to note that TGD people experience gender dysphoria at different levels – some may not experience any at all. Alongside this, avoidance of procedures vary depending on the individual. Levels of dysphoria may not directly correlate with whether a TGD person avoids a cancer screening procedure as causes are multifactorial.

On a broader scale, social stigma negatively impacts the health of TGD people through minority stress, alongside violence and victimisation. Factors known to be associated with cancer risk [17,18] are more common in TGD compared to CG people, potentially due to minority stress. As demonstrated in a UK-based study of 260,000 CG and 7,000 TGD participants, [19] transmasculine (TM) people had the highest prevalence of obesity (27.5%) as well as current and “ever smoking” (33.7% and 60.2%, respectively), while transfeminine (TF) people had the highest prevalence of dyslipidaemia (15.1%), diabetes (5.4%), hepatitis C (0.7%), and hepatitis B (0.4%). HIV infection was higher in TM and TF people (0.5% and 0.8%, respectively) compared to CG men (0.2%) and women (0.1%).

Furthermore, physical and sexual violence are unfortunately common experiences, [20] especially for transgender women, and “structural violence such as barriers to gender-affirming[care]” increase the risk of TGD people of developing physical and mental health disorders [21]. For reference, gender-affirming care refers to any interventions that help a TGD person transition to present congruently with their gender identity, which may commonly include hormone therapy and surgical procedures.

To determine whether cancer screening uptake in TGD populations is disparate, we performed the first systematic review analysing attendance rates for screening of all cancers with available data. We collated quantitative data on cancer screening attendance within TGD groups to build upon previously published qualitative reviews on the same topic. Some have addressed the gaps in the existing literature, noting the lack of culturally competent interventions to reduce healthcare disparities [22]. This review has the potential to quantitate the degree of inequity experienced by TGD patients from the current literature and provide insights from qualitative studies on how the inequalities created by our current healthcare systems could be addressed.

### Objectives

This is the first systematic review and meta-analysis to compare attendance rates for cancer screening between TGD and CG people. The primary aim is to determine whether there are differences in service utilisation and the secondary aim to investigate whether uptake changes based on the anatomical organ being screened.

### Language use

This review acknowledges that codifying gender identities into strict categories may overlook complexities surrounding the topic. Hence, we opted to use terms that encapsulate a broader range of identities while still maintaining structure for analysis.

We chose the terms “transmasculine” (TM), “transfeminine” (TF) and “gender non-conforming” (GNC) to categorise TGD identities in data extraction and analysis. In this scenario, we define TM as people who were assigned female at birth but identify with masculine identities. TF is defined as people assigned male at birth who identify with feminine identities. GNC includes people who do not strictly identify with either masculine or feminine identities. These decisions accommodate variations in language and reflect our current understanding of its influence on attitudes towards LGBTQ+ communities [23].

## METHODS

This systematic review adheres to PRISMA guidelines [24] and included the development of a protocol dataset[25] and prospective registration with PROSPERO 2022 CRD42022368911.

### Eligibility criteria

Eligible studies were required to be cross-sectional or case-control in design and had to include quantitative data relevant to the following PECO framework [26]dataset [25] –

- Participants: adults eligible for cancer screening services
- Exposure: TGD identity – whereby living as TGD within current societal systems may affect the outcome
- Comparator: CG identity – whereby gender congruence is the social norm so theoretically will not affect the outcome
- Outcome: attendance of cancer screening procedures in percentages or odds ratios

All types of cancer screening for different anatomical parts were included in the eligibility for the outcome component – this is for later comparison at the stage of synthesis. Studies without data or only qualitative data on the outcome of interest were excluded (Table 1). Further exclusions included data on cancer diagnosis either on symptomatic presentation or as incidental findings. There were no limits imposed on study settings due to paucity of available papers.

**Table 1.** Inclusion and exclusion criteria.

| Inclusion | Exclusion |
| --- | --- |
| Human studies | Animal studies |
| Adult participants (≥ 18 years old) | Participants under 18 years old or not eligible for cancer screening |
| Transgender or gender-diverse patients (include studies that also gather data from cisgender patients as reference) | Patient gender is not identified or only cisgender patients studied |
| Cross-sectional or case-control studies | Conference abstracts and posters, unpublished work, and review articles |
| Quantitative data on patient attendance of cancer screening services | Qualitative data |
| All cancer screening services offered to patients (e.g., cervical, breast, colon) | Cancer diagnosis from symptomatic presentation or unintentional findings |

Regarding report characteristics, there were no limits on the year of dissemination, but studies needed to be written in or translated to English. We excluded conference abstracts and posters, unpublished work, and review articles.

The main groups used in the synthesis will be the different anatomical parts for each type of cancer screening; the sub-groups will be broad TGD identities (i.e., TM, TF, and GNC) to allow inter-population comparison.

### Information sources

We conducted searches using four online databases (PubMed, EMBASE [via Ovid], CINAHL Complete [via EBSCO], and the Cochrane Library). A further “backward snowballing” step was used to extend the capture of literature to the systematic review [27]. This involved performing the screening process on citations identified within review articles that were excluded as part of the systematic review process.

### Search strategy

Development of the search strategy was based on index terms found in three to six sentinel articles that an initial PubMed screen of the literature identified. The full search strategy used the above PECO framework to provide structure for the search. Reviewers used the following MeSH terms and variations thereof: cancer, screening, transgender, and attendance. As per the eligibility criteria, we identified manuscripts from their inception until 30/09/2023 and did not set limits on language or location.

### Selection process

The screening process used Microsoft Excel, where search results were exported, and duplicates removed. Two reviewers (AC and CJ) screened articles based on title and abstract and both performed the backward snowballing step. Manuscripts chosen for further assessment were retrieved and read fully.

Reviewers followed the pre-specified inclusion and exclusion criteria but were blinded to each other’s decisions until screening was complete. Where there were differences in chosen articles, AC and CJ undertook discussions, each presenting the title and content of their articles and comparing in detail with the eligibility criteria. A third reviewer (BG) resolved any remaining disagreements.

### Risk of bias assessment

Quality assessment of the selected studies followed the Newcastle Ottawa Scale (NOS) for Assessing the Quality of Nonrandomized Studies in Meta-Analysis [28]. Assessed characteristics were selection, comparability, and outcome. The original NOS criteria were only specific to cohort and case-control studies, so these were modified to suit cross-sectional studies using existing publications. Papers received ratings of good, fair, or poor for risk of bias using previously published thresholds for converting NOS scores to Agency for Healthcare Research and Quality standards [29].

### Data collection process

All reviewers piloted a data extraction form on Microsoft Excel using several manuscripts. Extracted data included information such as publication information (i.e., title, authors, publication year, DOI, location), study type, type of cancer screening involved, number of participants involved, participant demographics, results on rates of attendance, and more. CJ independently extracted data from studies rated good to fair during risk of bias assessment and resolved any queries via open discussion with AC and BG. Where there was missing data, CJ reached out to relevant authors via email with variable responses.

### Data synthesis

We undertook meta-analysis and narrative synthesis using quantitative data on rates of cancer screening attendance among TGD participants. Measured outcomes included up-to-date attendance (UTD) and lifetime attendance (LT) of cancer screening involving varying anatomical locations.

UTD is the proportion of participants attending screening within recommended recall timeframes – this is dependent on the type of cancer screening and the guidelines used by each study as reference. Lifetime attendance (LT) refers to the proportion of participants having attended a certain screening service at least once in their life.

This data was collected as crude attendance rates in percentages and unadjusted odds ratios (OR). Adjusted OR were permitted for inclusion if the latter was not reported. The review presents the meta-analysed results as forest plots using unadjusted OR, representing the odds of a TGD person attending cancer screening in comparison to a CG person.

All articles with full text that were rated good or fair through the modified NOS were deemed eligible for synthesis. Those rated poor were excluded from data collection and analysis to reduce their influence on risk of bias. This choice was made as we recognised the greater probability of high risk of bias in our obtained studies – these were observational and often retrospective in nature. Prior to synthesis, the data were organised on Excel by screened organ, categorised into UTD or LT, and gender identity.

Estimates and their standard errors were entered directly into RevMan under the ‘generic inverse variance’ outcome. The software determined random-effects meta-analysis using the DerSimonian and Laird model [30], along with assessments of heterogeneity. Random-effects analysis was chosen because many variables differed between studies, e.g., within participant characteristics or study design. χ² and I² tests measured the presence and extent of heterogeneity, giving an estimate of how much studies varied. This allows some indication of how reliably results could be interpreted.

Meta-analysis used at least three studies per organ screened, as that was the minimum number of datasets available to us for each comparison. Syntheses were performed separately for UTD and LT data. The forest plots included subgroups by gender identity to visualise the distribution of results within TGD populations. We further synthesised the overall results for each organ group, subgrouping using UTD and LT status, to estimate the summary effect.

Narrative synthesis substituted meta-analysis for studies not suitable for the method of grouping used. For this same reason, we were unable to conduct Synthesis Without Meta-analysis [31] in lieu of traditional narrative synthesis [32] as originally planned. Any trends and relevant report findings noted by the authors after thorough appraisal of each paper were summarised.

## RESULTS

### Study selection, quality assessment, and study characteristics

The searches amassed 2425(AC)/1833(CJ) manuscripts, of which 277/208 were duplicates (Figure 1). The Protocol and Search terms for each database (Supplementary Data I) [dataset][25] were prospectively registered with PROSPERO [33]. The screening process (Supplementary Data II)[dataset] [25] identified 25 eligible records, whereby 18 met risk of bias and quality assessment requirements determined using NOS (Tables 2 and 3; Supplementary Data III) dataset [25].

**Figure 1.**
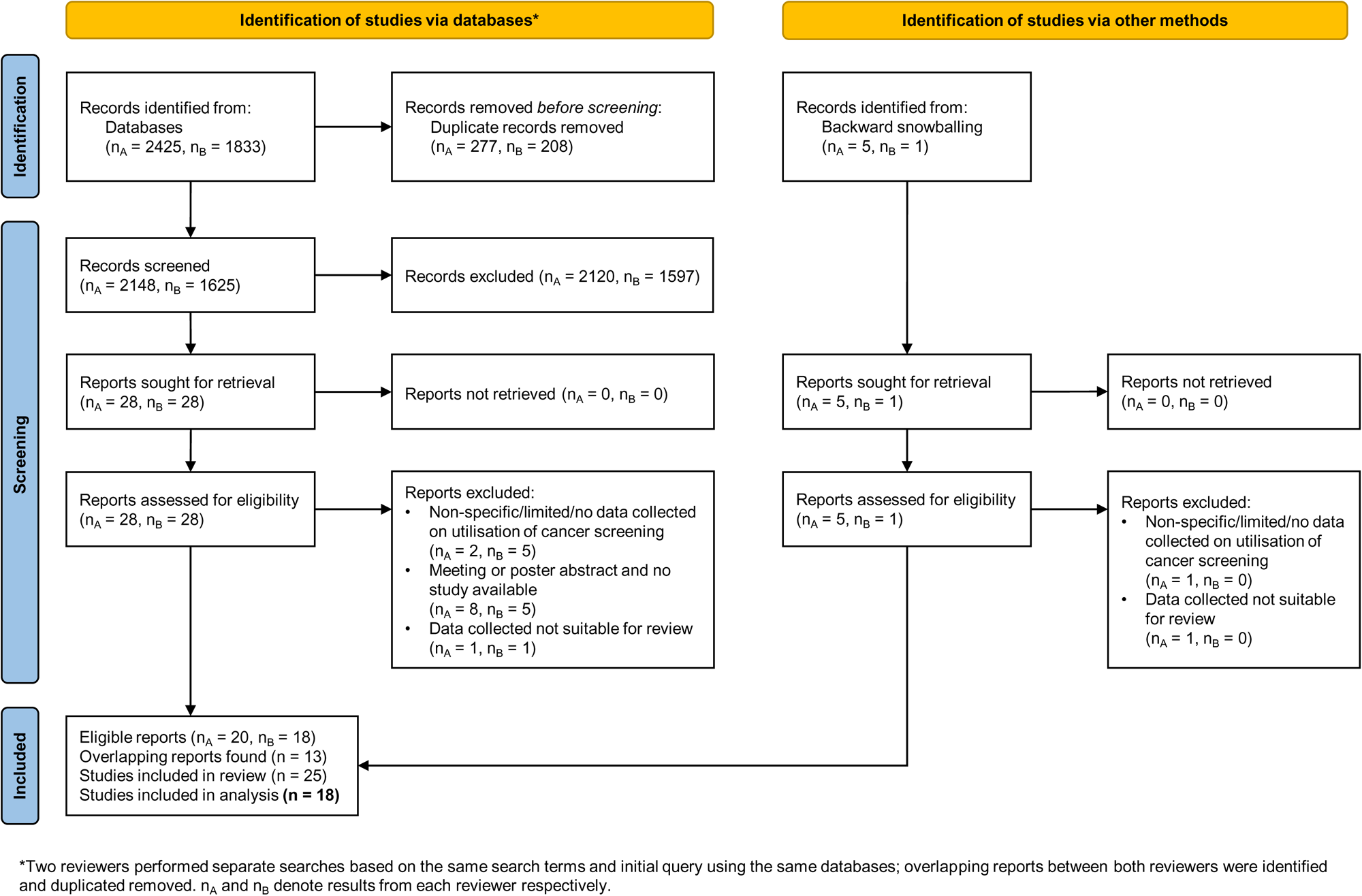
Prisma-P flow chart. Diagram showing the selection process of relevant articles, screened by title, abstract and full text, prior to quality assessment and meta-analysis.

**Table 2.**
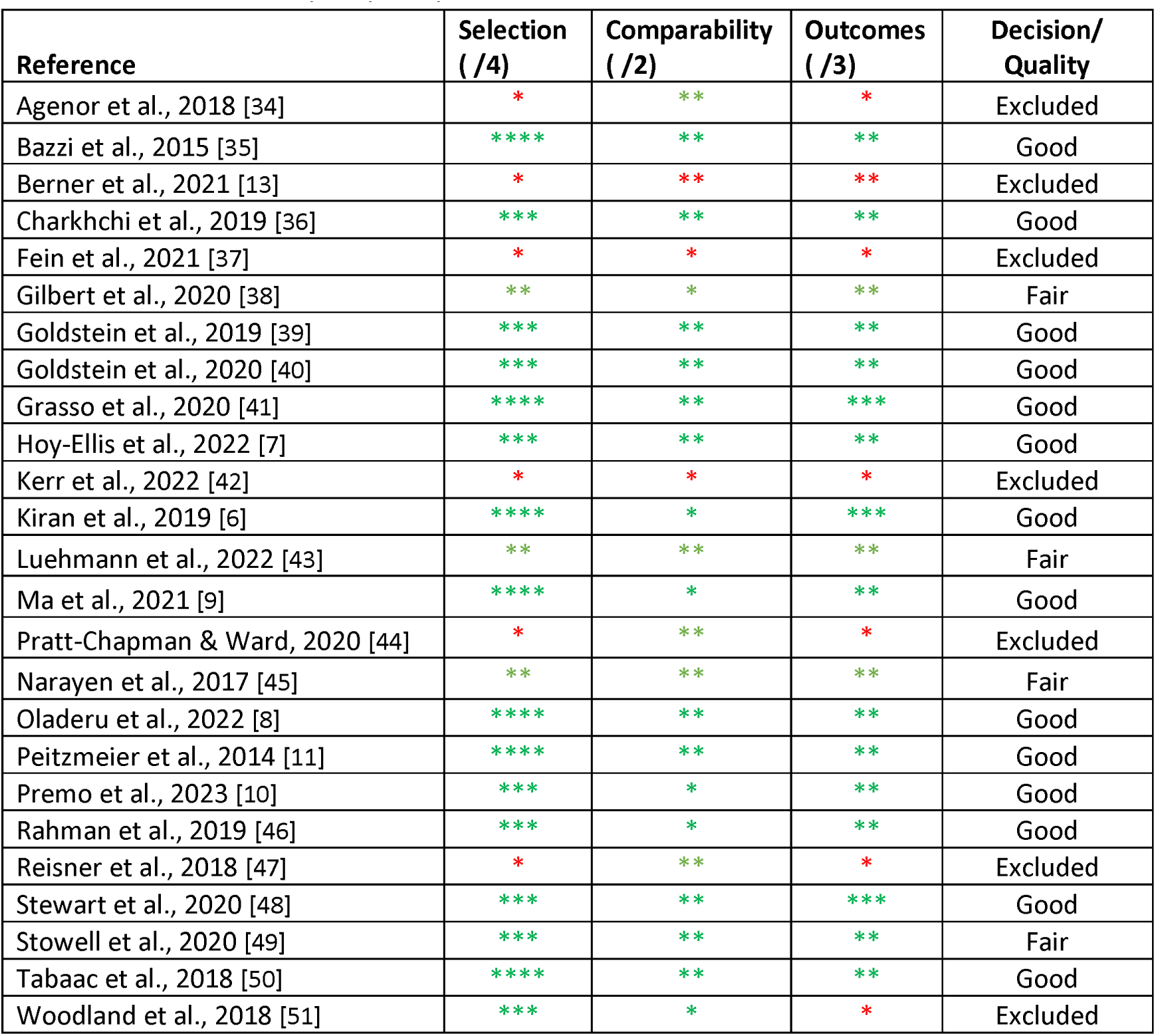
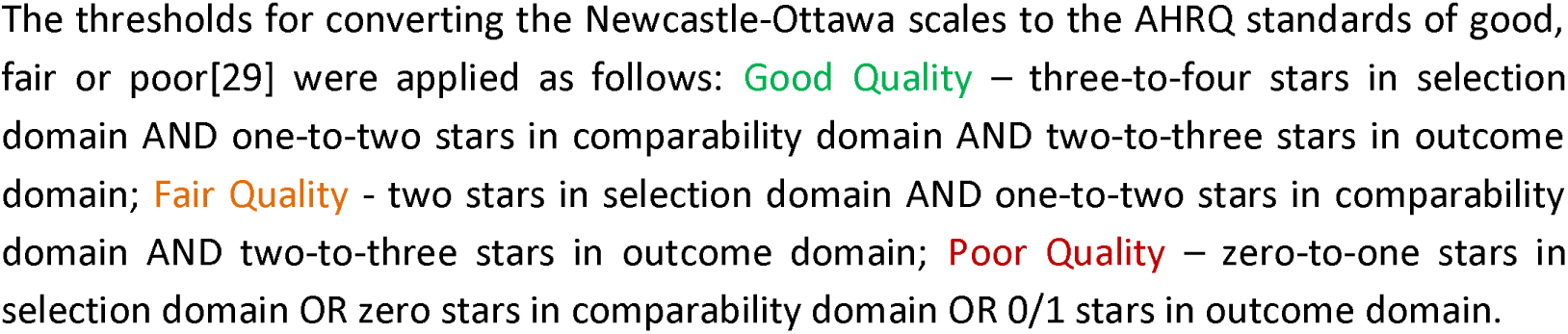
Risk of bias and quality analysis (NOS)

| Reference | Selection<br>( /4) | Comparability<br>( /2) | Outcomes<br>( /3) | Decision/<br>Quality |
| --- | --- | --- | --- | --- |
| Agenor et al., 2018 [34] | * | ** | * | Excluded |
| Bazzi et al., 2015 [35] | **** | ** | ** | Good |
| Berner et al., 2021 [13] | * | ** | ** | Excluded |
| Charkhchi et al., 2019 [36] | *** | ** | ** | Good |
| Fein et al., 2021 [37] | * | * | * | Excluded |
| Gilbert et al., 2020 [38] | ** | * | ** | Fair |
| Goldstein et al., 2019 [39] | *** | ** | ** | Good |
| Goldstein et al., 2020 [40] | *** | ** | ** | Good |
| Grasso et al., 2020 [41] | **** | ** | *** | Good |
| Hoy-Ellis et al., 2022 [7] | *** | ** | ** | Good |
| Kerr et al., 2022 [42] | * | * | * | Excluded |
| Kiran et al., 2019 [6] | **** | * | *** | Good |
| Luehmann et al., 2022 [43] | ** | ** | ** | Fair |
| Ma et al., 2021 [9] | **** | * | ** | Good |
| Pratt-Chapman & Ward, 2020 [44] | * | ** | * | Excluded |
| Narayan et al., 2017 [45] | ** | ** | ** | Fair |
| Oladeru et al., 2022 [8] | **** | ** | ** | Good |
| Peitzmeier et al., 2014 [11] | **** | ** | ** | Good |
| Premo et al., 2023 [10] | *** | * | ** | Good |
| Rahman et al., 2019 [46] | *** | * | ** | Good |
| Reisner et al., 2018 [47] | * | ** | * | Excluded |
| Stewart et al., 2020 [48] | *** | ** | *** | Good |
| Stowell et al., 2020 [49] | *** | ** | ** | Fair |
| Tabaac et al., 2018 [50] | **** | ** | ** | Good |
| Woodland et al., 2018 [51] | *** | * | * | Excluded |

The thresholds for converting the Newcastle-Ottawa scales to the AHRQ standards of good, fair or poor[29] were applied as follows: **Good Quality** – three-to-four stars in selection domain AND one-to-two stars in comparability domain AND two-to-three stars in outcome domain; **Fair Quality** - two stars in selection domain AND one-to-two stars in comparability domain AND two-to-three stars in outcome domain; **Poor Quality** – zero-to-one stars in selection domain OR zero stars in comparability domain OR 0/1 stars in outcome domain.

**Table 3.**
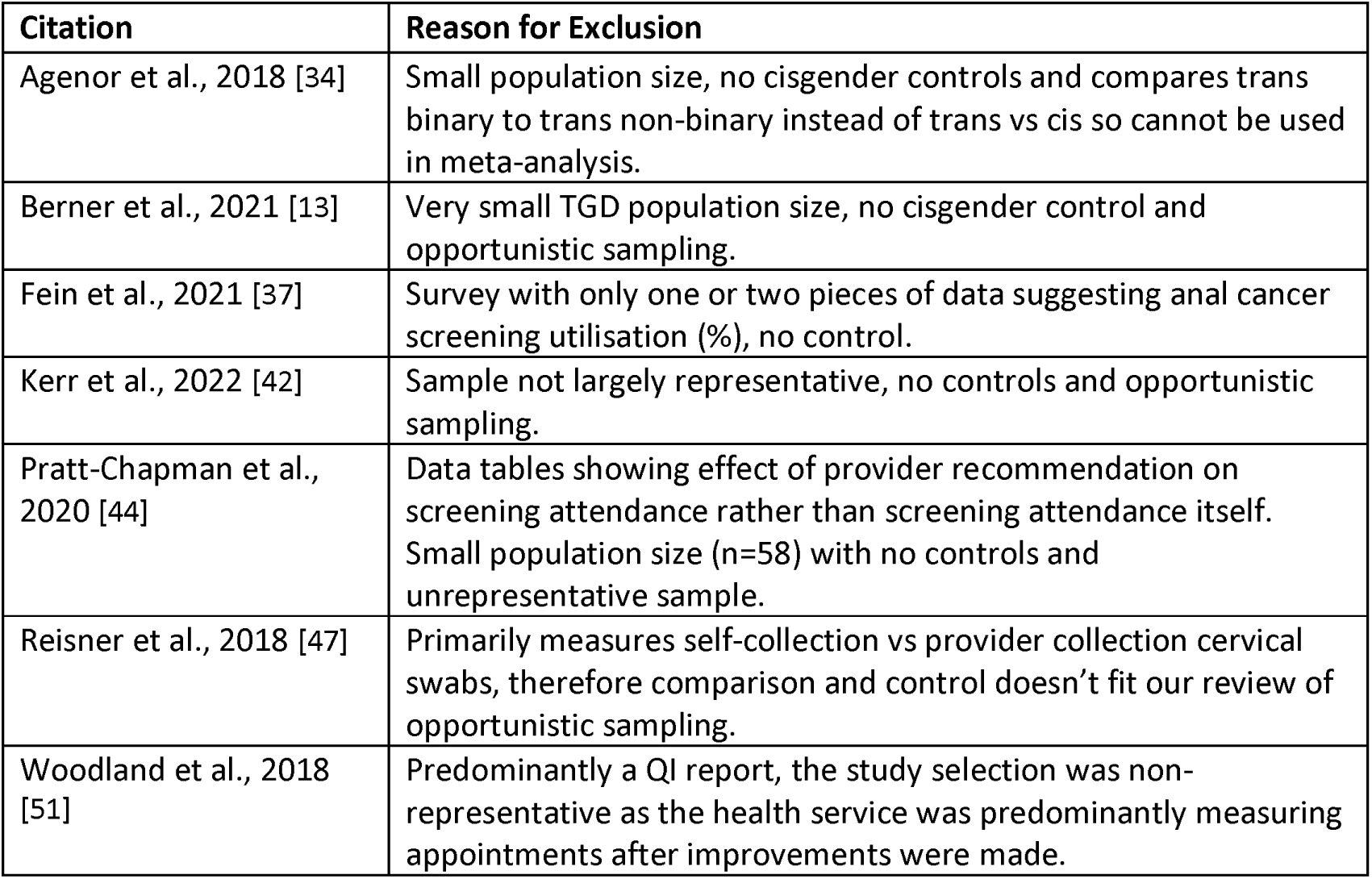
Articles removed following NOS and reasons for exclusion.

17 selected papers were from the US and one was from Canada. Publication years ranged from 2015 to 2023. Papers accepted for data extraction were cross-sectional studies, including retrospective chart reviews and survey analyses (Supplementary Data IV)[dataset] [25]. The data represented cancer screening for four different anatomical parts. Eight studies described breast cancer [6–8,35,36,38,48,50], ten described cervical cancer [6–8,11,36,39,40,46,48,50], four described prostate cancer [7,9,10,50] and three described colorectal cancer [6,36,50]. Six articles reported results on multiple organs [6–8,36,48,50], therefore increasing the pool of data available for analysis.

Meta-analysis for UTD versus LT was performed separately for each of the four cancer screening categories to maximise data capture – i.e., breast, cervix, prostate, colorectal. 11 of 18 studies were added to our meta-analysis [6–11,35,36,39,46,50]. Seven of 18 studies [38,40,41,43,45,48,49] could not be included in the quantitative synthesis due to missing data or lack of control groups prohibiting the calculation of OR. For instance, some papers did not have confidence intervals, and some used secondary data from national censuses for their CG comparators. We contacted the corresponding authors of these articles for additional information or raw data but did not receive the necessary details required for meta-analysis.

### Up-to-date attendance

Meta-analysis identified that the discrepancies for UTD cervical screening (Figure 2Ai) in TGD people were OR=0.37 (95% CI [0.23, 0.60], p<0.0001). There was no TF subgroup due to the requirement of a uterine cervix. UTD mammography screening (Figure 2Bi) also showed discrepancies overall (OR=0.41, 95% CI [0.20, 0.87], p=0.02). UTD results for prostate and colorectal screening were not meaningfully different to CG attendance (Figure 2Ci, 2Di).

**Figure 2.**
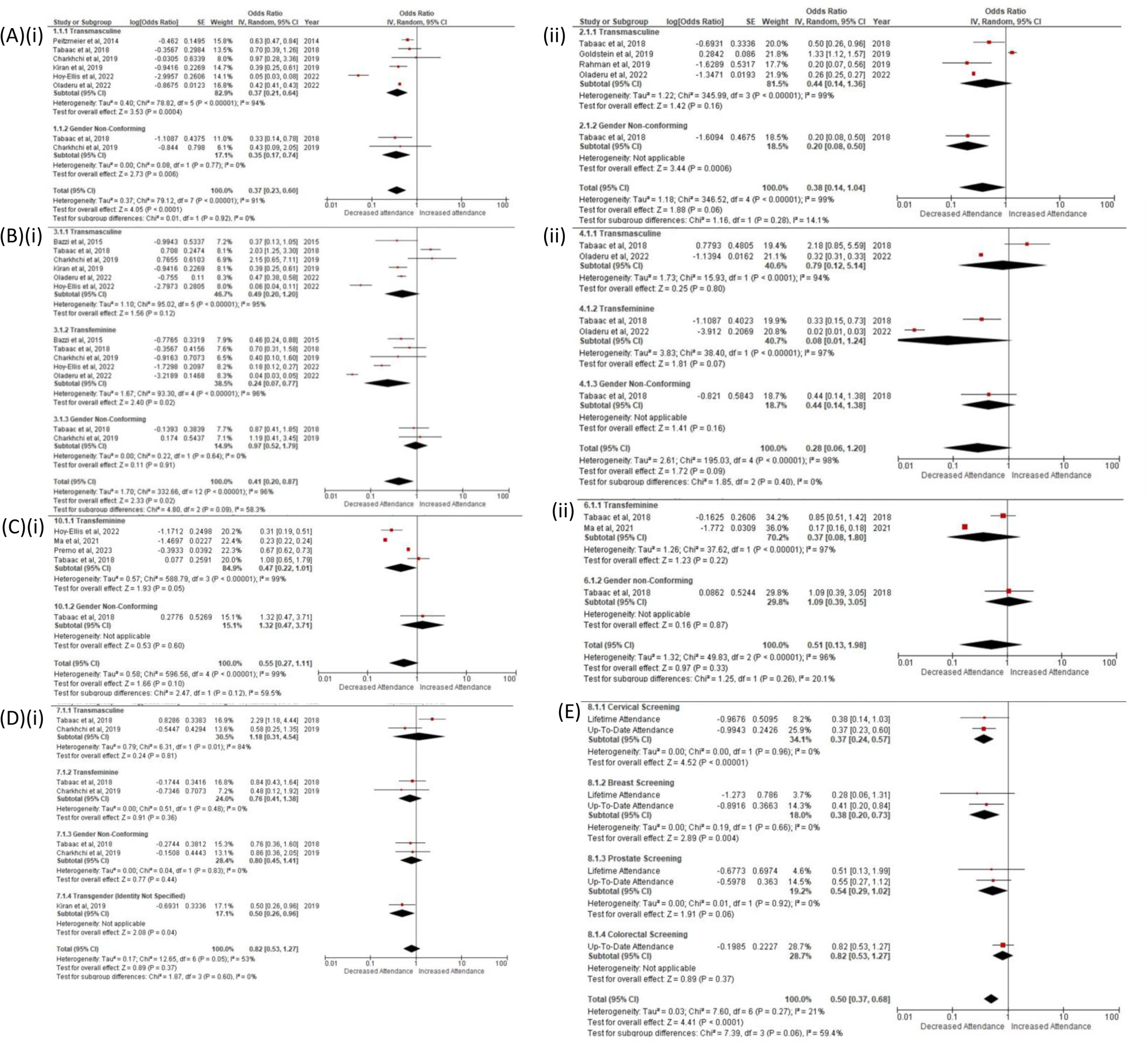
Forest plots for the meta-analysis of TGD individuals at cancer screening appointments. Random-effects meta-analysis shows up-to-date (i) and lifetime attendance (ii) of TGD patients for (A) Cervical; (B) Breast; (C) Prostate (D) Colorectal and (E) all cancer screening.

Common findings from studies not included in meta-analysis generally supported our results. TM individuals were less likely to be up to date with cervical screening [40,41,48]. UTD rates for mammography screening were also reduced for TGD people, with no differences between TGD populations [43,48].

In contrast, one paper found comparable attendance of mammography screening between TGD and CG participants within the most recent two years when analysing data from the 2014 Behavioural Risk Factor Surveillance System (BRFSS) survey [45].

### Lifetime attendance

All LT results in meta-analysis found similar rates of attendance between TGD and CG data (Figure 2Aii, 2Bii, 2Cii). There was no LT data available for colorectal cancer screening. In narrative synthesis, no difference was found in LT rates for cervical screening between TGD and CG participants [38], but lower LT rates were reported for mammography screening [38,43].

### Summary effect

Overall, data synthesis displayed a trend of reduction in both UTD and LT for TGD people compared to CG people, but the separate analyses only found differences for UTD cervical and mammography screening. However, pooling total OR for all LT and UTD syntheses showed reduced attendance in the TGD group altogether (OR=0.50, 95% CI [0.37, 0.68], p<0.0001; Figure 2E).

We noted that despite having less data for colorectal screening (i.e., no LT data), results trended towards showing less disparity when compared to other cancer screening.

Findings from separately analysing results from papers ineligible for meta-analysis also predominantly supported our syntheses.

### Lung cancer screening

We found one study investigating lung cancer screening via analysis of the 2017 and 2018 BRFSS surveys. This was not a screening type included in the meta-analysis because unlike breast, cervical, or bowel cancer, very few countries have national screening programmes for lung cancer. For instance, the UK only announced the rollout of a targeted lung cancer screening programme this year [52]. The authors found that despite similar eligibility and smoking statuses within their TGD and CG groups, the former attended less than the latter at 2.3% and 17.2%, respectively [49].

## DISCUSSION

Overall, TGD individuals are less likely to utilise breast and cervical cancer screening than their CG counterparts, with no meaningful differences found in prostate and colorectal screening. The biggest disparity in attendance was seen specifically in UTD cervical screening.

Levels of discomfort and invasiveness could contribute to this distribution of results. Cervical screening uptake rates in CG women remain low worldwide, likely due to difficulties tolerating examination [53]. This effect is compounded by multiple factors in TGD people. For example, androgen therapy has been associated with increased odds of failure to obtain adequate cervical cytology samples [54] and increased technical difficulty in examination due to atrophic changes to vaginal and cervical tissue [55]. This may necessitate repeated examinations or cause increased discomfort, pain, and gender dysphoria, contributing to avoidance of cervical screening [54].

More research is required to investigate differences between TGD identities. One study found higher rates of healthcare avoidance caused by anticipated discrimination in transgender men compared to transgender women (adjusted odds ratio [AOR] = 1.32, 95% CI [1.21, 1.45]) [36]. Non-binary and genderqueer individuals in this study were reported to avoid healthcare less (AOR = 0.71, 95% CI [0.63, 0.80]) but experience more misunderstanding from providers and make more effort to conceal their TGD identity [15].

### Limitations

This review analysed cross-sectional studies and required modification of the NOS criteria, deviating from the validated framework of the original scoring system. Despite excluding studies determined to be “poor” for risk of bias, most included in our analysis had a higher risk of bias due to their observational or retrospective nature.

Most included studies also analysed survey data, likely introducing volunteer bias. For example, three studies included in the meta-analysis used data from the BRFSS [8,36,50], and two of these include data from the 2016 BRFSS.

This review was limited by the paucity of available data. The search process could have been more comprehensive by including results from other literature like meeting abstracts. The process also required greater standardisation in implementation, i.e., identical search strategies between authors.

We lacked the resources required to perform further statistical tests for detailed analysis of the results. For example, meta-analysis subgroups were unpowered, so this data could not be reliably interpreted. Statistical heterogeneity was high, where I > 90% for most analyses. This was likely attributable to broad inclusion criteria in searches and differences in study design but could not be reliably investigated.

### Implications

TGD people may be less likely to attend cancer screening due to multiple structural and individual determinants. For example, socioeconomic status independently impacts cancer screening rates through income, insurance and healthcare access. TGD people are more likely to have lower incomes and higher rates of unemployment [36], affecting screening uptake. Prioritisation of basic needs may take precedence over accessing cancer screening in people with financial or housing insecurity [56].

Health insurance, a major consideration in countries like the USA, can present barriers as well. Cancer screening may not be free on every insurance plan or may be inaccessible to those without insurance [57,58].

Education may affect engagement with cancer prevention services. While emotional distress strongly prevents LGBTQ+ people from accessing cancer screening, advanced education and increased age are correlated with lower levels of emotional distress [59]. Better patient education could improve screening rates. Several studies suggest that TGD people may receive less education about HPV and its links to head, neck and oral cancers than CG people [46,59], but would be just as likely to understand these risks once educated. Compounded with greater vulnerability to distress surrounding cancer screening and healthcare, TGD people are likely more at risk of poor adherence with screening for HPV-related cancers.

Causes of emotional distress around cancer screening can include anxiety due to anticipated or experienced discrimination [15] and gender dysphoria from intimate procedures or heavily gendered healthcare environments [5]. Poor education of healthcare providers (HCPs) and administrative staff contributes to prejudice against TGD individuals, ranging from avoidance of conversations about screening and safe sex practices to outright discrimination through refusal to provide adequate care [13].

Evidently, a greater number of studies investigating the experiences of TGD people all across the world is required to better explore what disparities TGD people experience in cancer screening and what factors affect degrees of disparity. Gaps in data exist regarding differences experienced by GNC people. New studies may consider including participant characteristics specific to TGD populations, such as the type of transition being undertaken (e.g., medical, social, none) and the length of time that has been involved. Reasons for inter-group differences would benefit from intersectional analyses to evaluate how racism, classism, and sexism may affect this in cancer screening.

Further research into how hormone therapy affects susceptibility to cancers and whether this affects how clinicians should approach cancer screening guidelines for TGD populations is needed. For instance, eligibility for breast screening in TF individuals may need to be guided by current age and length of time exposed to feminising hormones [60].

Improved patient engagement with cancer screening may be facilitated by designing healthcare structures for better accessibility [59], e.g., providing gender-neutral environments in settings like breast screening clinics, online systems for scheduling appointments [40,61], improving medical coding for TGD identities, and implementing inclusive cancer screening protocols. Other actions include considering more acceptable alternatives like self-collected HPV swabs [40,61], formal training of HCPs to tackle ignorance and discrimination [62], targeted patient education specific to cancer screening and TGD identity with inclusive language [63], and consideration of an “organ-based approach” whereby screening recall is based on the relevant organs present [64].

The relationship between TGD status and cancer screening requirements can be poorly understood by both patients and HCPs, leading to deviations from recommended guidelines as seen in cervical screening [65,66]. Correlations between lower socioeconomic status and lower cancer screening rates suggest that interventions like HCP or patient education alone will not suffice in reducing disparities.

### Conclusions

While this systematic review supports the hypothesis that TGD people have lower rates of accessing cancer screening, suggestions of areas for improvement are inferred and not conclusive. Historically, interventions implemented to address health care disparities experienced by TGD patients have lacked cultural competence [22]; it is imperative to consider this when exploring new interventions.

There are still individual and institutional barriers preventing TGD people from accessing cancer screening services. Further investigations would provide insight into the degree of inequity experienced by TGD patients. Bridging this gap will require consolidated efforts from healthcare systems with a “multi-level and multi-faceted approach” [15]. The joint production of future interventions with the TGD community is vital to improving both cancer screening experience and outcomes. Good examples include accessible guides devised with TGD individuals, self-sampling programmes, and targeted screening programmes.

## Data Availability

All data produced are available online at https://doi.org/10.5281/zenodo.10621705

## Acknowledgements

We would like to thank Dr Heidi Baseler who runs the INSPIRE program at the Hull York Medical School, Danny Fletcher for assistance with figures and all patients whose data contributed to this work.

## Ethics Approval Statement

Research ethics committee or institutional review board approval was not required for this systematic review but only articles which were peer-reviewed and reasonably believed to have received research ethics approval were included.

## Contributor and Guarantor Information

AC had the idea for the article, AC and CJ performed the literature search, AC, CJ and BG wrote the article. The planning, conduct, and reporting of the work described in the article was performed by AC, CJ and BG. HD and SO’C provided unique insights based on lived experience while HD, BG and SO’C provided specialist knowledge into the topics discussed.

The corresponding author (BG) attests that all listed authors meet authorship criteria and that no others meeting the criteria have been omitted.

The guarantor (BG) accepts full responsibility for the work and/or the conduct of the study, had access to the data, and controlled the decision to publish.

## Patient and Public Involvement statement

It was not appropriate or possible to involve patients or the public in the design, or conduct, or reporting, or dissemination plans of our research.

## Copyright/licence for publication

The Corresponding Author has the right to grant on behalf of all authors and does grant on behalf of all authors, worldwide licence to the Publishers and its licensees in perpetuity, in all forms, formats and media (whether known now or created in the future), to i) publish, reproduce, distribute, display and store the Contribution, ii) translate the Contribution into other languages, create adaptations, reprints, include within collections and create summaries, extracts and/or, abstracts of the Contribution, iii) create any other derivative work(s) based on the Contribution, iv) to exploit all subsidiary rights in the Contribution, v) the inclusion of electronic links from the Contribution to third party material where-ever it may be located; and, vi) licence any third party to do any or all of the above.

## Abbreviations

AOR: adjusted odds ratio
BRFSS: Behavioural Risk Factor Surveillance System
CG: cisgender
CI: confidence interval
GNC: gender non-conforming
HPV: Human Papilloma Virus
LT: lifetime
NOS: Newcastle-Ottawa scale
OR: odds ratio
TF: transfeminine
TGD: transgender and gender-diverse
TM: transmasculine
UTD: up-to-date

